# Time to sustained control of the effective reproduction number as a failure-time estimand: a county-level analysis of the first SARS-CoV-2 wave in the United States

**DOI:** 10.64898/2026.11.05.26367273

**Authors:** Athanasios Kakasis, Marianna Panagiotidou, Nikolaos Demiris, Evangelia Samoli, Vana Sypsa

**Affiliations:** Department of Preventive Medicine, Epidemiology and Medical Statistics, School of Medicine, National and Kapodistrian University of Athens, Greece; AINIGMA Technologies BV, Leuven, Belgium; Department of Statistics, Athens University of Economics and Business, 76 Patission Str., 10434 Athens, Greece

**Keywords:** SARS-CoV-2, COVID-19, reproduction number, survival analysis, Cox model, county-level, United States, policy stringency, population density

## Abstract

Subnational analyses of emerging pathogen transmission typically model the reproduction number (R_t_) as a continuous outcome and estimate average associations with its level. Operationally, however, response planning depends on a different quantity: which jurisdictions bring transmission below the epidemic threshold (R_t_<1), and how quickly. We formulate time to control as a failure-time estimand and show that doing so exposes a structural feature that linear regression does not accommodate: a jurisdiction cannot enter the risk set until enough cases accumulate to estimate R_t_. Cases accumulate faster where transmission is faster, so entry time depends on the outcome process itself and on factors that drive early spread. We account for this outcome-dependent observability using a mixed-effects Cox proportional hazards model with calendar-time left truncation and state-level Gaussian frailties, applied to 2,956 US counties observed between January 22 and July 15, 2020. The primary event was defined as the first three-day sustained reduction of mean R_t_ below one. Control was rare (171 counties, 5.8%). Each standard-deviation increase in log population density was associated with a lower rate of control (hazard ratio 0.54; 95% confidence interval 0.44–0.67) and higher policy stringency with a faster rate (HR 1.50; CI 1.09–2.05). Evaluating jurisdictions by their rate of reaching an operational target - rather than by average transmission level - provides a clearer, decision-relevant metric for prospective allocation decisions and public health interventions.

## Introduction

When an emerging respiratory pathogen begins to spread within a country, the question of which subnational jurisdictions first reduce transmission below the epidemic threshold, and how quickly, is directly relevant to outbreak surveillance and resource planning. The instantaneous (effective) reproduction number R_t_ has become the standard real-time summary of transmission intensity at national and sub-national scale, with values below one indicating that local case incidence will, on average, decline(1). During the first wave of the COVID-19 pandemic, US states reported considerable heterogeneity in R_t_ across constituent counties. Still, the structural and policy correlates of this heterogeneity, and of the speed with which counties reached R_t_ < 1, remain incompletely characterized.

Previous studies have examined associations between county-level factors and R_t_ using regression models that treat R_t_ as a continuous outcome at each observation period(2) or by descriptively characterizing R_t_ trajectories with policy annotations(3–5). These methods treat each county-day as an observation and yield average effects on R_t_, marginalizing across calendar time. While suitable for estimating the impact of e.g. a 10-point increase in stringency on R_t_, they are less effective for assessing how long it takes a county with specific characteristics to achieve transmission control or for identifying which counties succeed.

An alternative approach is to define a binary event, specifically a sustained reduction of mean R_t_ below one, and to study the time from the first day of estimable R_t_ to the occurrence of this event using survival analysis. This framework produces hazard ratios that directly quantify differences in the rate of achieving control across counties with varying characteristics, while accounting for censoring of counties that do not achieve sustained control during the study period and for time-varying exposures such as policy stringency and temperature. Additionally, this approach clarifies an aspect that regression on R_t_ levels may obscure: counties enter the analytic risk set at different calendar dates, depending on when their cumulative case counts permit reliable R_t_ estimation, necessitating appropriate handling of differential entry to avoid bias.

Framing control as a time-to-event outcome exposes a structural feature that regression on R_t_ levels does not accommodate: a county becomes observable to the analysis only once case accrual permits R_t_ estimation, so its entry into the risk set is determined by the outcome-generating process. Entry is correlated with population density in particular because cases accumulate fastest where contact rates are highest, so dense counties become observable earlier than sparse ones. To our knowledge, sub-national transmission control has not previously been analysed as a left-truncated failure-time process in which risk-set entry is governed by outcome estimability. A related idea appears in the stochastic-process literature, where the time for R_t_ to first cross a threshold is derived analytically for a single epidemic trajectory and fitted to regional surveillance data(6). That approach characterizes when one epidemic process crosses a threshold, whereas the present analysis treats jurisdictions as units of observation and estimates covariate associations with the crossing hazard under delayed entry. This is not a new estimator but rather a new correspondence between an established survival-analytic structure and a recurring feature of surveillance data, in which an outcome is measurable only after data have accrued.

This study applied a mixed-effects Cox model with a calendar-time clock and left truncation to examine time to sustained control across the contiguous United States during the first wave of the COVID-19 pandemic (January-July 2020). The analysis evaluated the roles of population density, policy stringency, and temperature, and quantified residual state-level heterogeneity using a Gaussian frailty term. The objectives were to demonstrate the survival analysis framework as a methodological complement to regression-on-R_t_ approaches and to characterize both the counties that achieved sustained transmission control and the speed at which they did so. From a surveillance perspective, it is critical to understand not only the average reduction in transmission due to an exposure, but also how quickly transmission falls below the epidemic threshold and which jurisdictions do so. This information is essential for allocating limited early-response resources, such as testing, contact tracing, and surge support, during the initial weeks of a novel pathogen wave.

## Methods

### Data sources

County-level daily reported case counts for January 22 to July 15, 2020 were obtained from the Johns Hopkins University Coronavirus Resource Center(7). Population estimates and population per square mile were sourced from the US Census Bureau Population Estimates Program (2020 baseline)(8). Daily state-level policy stringency was obtained from the Oxford COVID-19 Government Response Tracker, using the weighted-average Stringency Index(9). Monthly mean temperature for each county from January through July 2020 was obtained from the NOAA National Centers for Environmental Information (NCEI)(10).

The analytic dataset comprised 2,956 counties for which we could estimate R_t_ at some point during the study window. We excluded an additional 150 counties that did not accumulate sufficient cases to support R_t_ estimation. The complete dataset used in this study is openly available through a research repository(11).

### Estimation of the effective reproduction number R_t_

For each county, we estimated mean daily R_t_ using the Cori et al. method implemented in the R package EpiEstim(12). We applied a parametric serial-interval distribution with a mean of 5.2 days and a standard deviation of 2.8 days(13). We used a seven-day sliding window and required at least 20 cumulative cases within a window before reporting R_t_. We smoothed daily incidence with a seven-day right-aligned moving average to account for weekday reporting effects.

### Event definition

The event of interest was sustained transmission control, defined as the first day of the first three-day period during which mean R_t_ was below one. We chose the three-day criterion over a single-day threshold to filter out brief stochastic crossings due to posterior uncertainty in R_t_. In our data, 91% of counties with any single day of R_t_ < 1 also sustained the crossing for at least three consecutive days, indicating the criterion preserves real events. Sensitivity to this choice was assessed using one-day and five-day sustained-crossing definitions.

### Basic reproduction number

For each county, we defined the basic reproduction number R_0_ as the first estimable mean R_t_, evaluated on the county’s entry date into the risk set. Because entry is defined as the first day on which R_t_ is estimable, this parameter is measured exactly at the left-truncation point and cannot incorporate post-entry information. Strictly, R_0_ denotes the basic reproduction number in a fully susceptible population, whereas this quantity is the effective reproduction number at the county’s first estimable day; we retain the label because entry occurs early in each county’s epidemic, when depletion of susceptibles is negligible.

### Survival framework

We used calendar time as the analysis clock, with each county entering the risk set on the first day R_t_ could be estimated (left truncation) and exiting at either the event date or July 15, 2020 (administrative censoring). Because a county R_t_ becomes estimable only after reaching the minimum case threshold, the entry date correlates with size and population density: denser, larger counties enter the observation earlier than sparse rural ones. The calendar-time clock with left truncation ensures each county is absent from risk sets before its entry date and is not considered to have failed to control before it was observable. Each county-day during follow-up contributed one row to a counting-process dataset (entry, exit, event-indicator), with time-varying covariates assigned the value observed on each day.

The primary model was a mixed-effects Cox model fit using the coxme package in R(15):

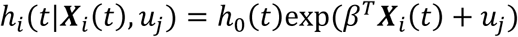

where i=1,.,2956 denotes the county; j=1,..,49 the state; ***X****_i_* is a vector of county-specific covariates comprising the variables *R*_0_ and population density (both log-transformed), alongside the time-varying variables stringency and temperature; u_j_ ∼ N(0, σ²) is a state-level Gaussian frailty representing residual baseline-hazard heterogeneity across 48 contiguous states and the District of Columbia. We entered R_0_ on the logarithmic scale. Under the assumption that behavioral and policy responses reduce transmission approximately multiplicatively(14), the reduction a county must achieve to reach the epidemic threshold is log R_0_ − log 1 = log R_0_, so log R_0_ is the distance to be traveled and is the quantity the hypothesis concerns; the natural scale was retained as a sensitivity analysis. We modeled stringency as a 14-day right-aligned rolling mean of the daily weighted Oxford Stringency Index, reflecting the lag between policy implementation and observable effects on transmission. We set temperature as a time-varying covariate equal to the mean calendar-month temperature for each county. We standardized all continuous covariates to have a mean of zero and unit variance. Population density was log-transformed before standardization. Because the baseline hazard is unspecified in calendar time, any signal shared across counties on a given date, including national seasonal warming, is absorbed into h₀(t).

### Estimand and interpretation

We distinguish the target of each coefficient. The population density and temperature coefficients are descriptive, predictive contrasts in the rate of reaching sustained control, not causal effects. We interpret the stringency coefficient as an association conditional on state-level frailty, which adjusts for shared state-level determinants (governance, health-system capacity, mobility); the early-outbreak-to-policy pathway remains as reverse causation that conditioning cannot remove (Figure 1). The same graph shows an open path from population density to the event through case ascertainment, which adjustment cannot close and which we address by quantitative bias analysis instead. We also adjust for the baseline reproduction number at entry as a proxy for early transmission intensity; because it is a function of observed case counts, conditioning on it could in principle open that same path, so we report the population density association with and without it.

**Figure 1:**
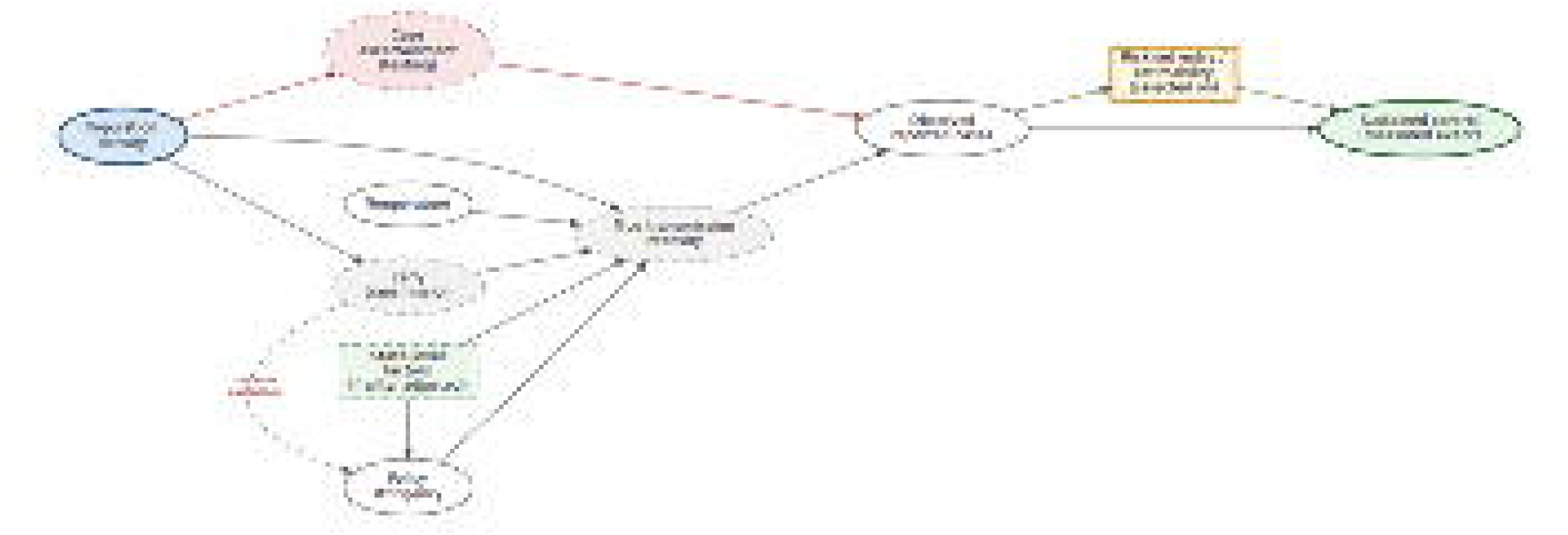
Assumed structural relationships (directed acyclic graph) for the population density-control analysis. Blue denotes the exposure (population density) and green the measured outcome (sustained transmission control); dashed ellipses are unobserved variables. The state frailty adjusts for shared state-level determinants (box), while the early-transmission-to-stringency edge represents reverse causation that adjustment cannot remove. Case ascertainment lies on an open path from population density to the observed event and cannot be closed by conditioning, which motivates the quantitative bias analysis; risk-set entry,(box) is the left-truncation selection node.

### Diagnostics

Proportional hazards (PH) were assessed using scaled Schoenfeld residuals(16). We assessed functional-form linearity using likelihood-ratio tests against models with natural cubic splines (3 knots at the 5th, 50th, and 95th percentiles), with a polynomial-deviation test (quadratic + cubic terms) as a complementary check. Influence was assessed via dfbeta against the Belsley threshold (2/√events)(17). Outliers were assessed via deviance residuals. Harrell’s C-statistic summarized model discrimination ability with 95% confidence intervals from a county-level cluster bootstrap (500 resamples).

### Sensitivity analyses

The primary specification was assessed for robustness using five sensitivity analyses: (i) refitting under one-day (188 events) and five-day (132 events) sustained-crossing definitions; (ii) restricting follow-up to calendar day 60 onward, addressing a marginal PH violation for stringency attributed to early-pandemic confounding; (iii) refitting with natural cubic splines on each continuous covariate to confirm the linear-in-log-hazard specification; (iv) entering the baseline reproduction number on the natural rather than logarithmic scale; and (v) allowing the coefficient for log R_0_ to differ before and after calendar day 60, testing the non-proportional effect implied by a distance-to-threshold mechanism.

Because a county enters the risk set only once its R_t_ becomes estimable, and estimability depends on case accumulation and therefore on population density, the way time at risk is handled can itself bias the population density association. To quantify what is lost by simplifying the time structure, we refitted the model and re-estimated the population density association under alternative specifications. Within the counting-process layout, the left-truncated fit was compared with one in which every county was treated as at risk from the calendar day zero and its earliest observed covariates were carried backward to that origin. Within a collapsed single-record cohort, in which each county contributes one row with time-averaged covariates, the left-truncated fit was compared with two variants: one treating all counties as at risk from calendar day zero (introducing immortal time bias) and one using a gap-time clock measuring time elapsed since county-specific entry. Each comparison is made within its own layout, so that the contrast isolates the handling of time rather than the cost of collapsing time-varying covariates.

### Quantitative bias analysis for case ascertainment

As reported cases reflect both underlying infection dynamics and local testing capacity, observed county-level associations with population density may be confounded by differential case ascertainment. The Cori estimator calculates R_t_ as the ratio of current incidence to a serial-interval-weighted sum of recent incidence, so a reporting fraction that is constant over time cancels out. Therefore, R_t_ estimates are biased by time-varying ascertainment rates. To model this potential bias, we assumed that testing expanded smoothly over calendar time, possibly faster in denser counties, with a single parameter γ which dictates how much faster testing expanded in high-density relative to low-density counties (γ = 0: testing expanded at the same rate across all counties regardless of population density). Rather than re-estimating R_t_ from case counts adjusted for ascertainment, we rescaled each county’s existing R_t_ series analytically using the renewal structure of the estimator. This transformation reproduces the primary series exactly when γ = 0. From each rescaled series across a grid of γ, we re-derived the sustained-control event and refitted the mixed-effects Cox model, tracing the hazard ratio for population density. Rescaling does not change which county-days are estimable, so risk-set entry is unchanged, and only the events are shifted. Parameter γ is a sensitivity parameter evaluated across a range of plausible assumed values rather than an empirically estimated quantity. The analysis does not address changes in the serial interval or in who was tested.

### Software

All analyses were performed in R 4.5.2(18). Specific packages: EpiEstim for R_t_ estimation; survival and coxme for survival modeling; zoo for rolling-window covariate construction; dplyr and tidyr for data manipulation.

## Results

### Cohort and event description

The analytic cohort included 2,956 US counties, followed for a median of 106 days (IQR 90–114) among counties that did not achieve the event and 41 days (IQR 22–64) among those that did (Table 1). Sustained transmission control was uncommon: 171 counties (5.8%) achieved the event during the study window. Counties achieving the event were generally smaller (median population 16.8 vs 29.1 thousand) and less dense (35.9 vs 49.4 persons per square mile) than those that did not, consistent with population density as a dominant structural correlate of time to control. Mean stringency over follow-up was higher among event counties (weighted index 64.5 vs 57.4). Latitude and January–July mean temperature were similar between groups (37.9 vs 38.2; 56.0 vs 55.7 °F, respectively). Regional composition differed modestly, with the Northeast underrepresented among event counties (2.9% vs 7.5%) and the Midwest overrepresented (40.9% vs 33.0%). April 2020 was the most common month of risk-set entry among event counties (67.3%) and March 2020 among counties not achieving the event (30.3%).

**Table 1.** Baseline characteristics of US counties achieving and not achieving the event of sustained COVID-19 transmission control (mean R_t_ < 1 for ≥ 3 consecutive days) during January 22 – July 15, 2020. Continuous variables presented as median (IQR); categorical variables as n (%). Wilcoxon rank-sum tests for continuous variables; chi-square tests for categorical variables.

| Variable | Event (n = 171) | No event (n = 2,785) | p-value |
|---|---|---|---|
| Population (thousands) | 16.8 (8.7, 31.2) | 29.1 (13.0, 78.8) | <0.001 |
| Population density (persons/sq mi) | 35.9 (15.1, 62.4) | 49.4 (20.4, 138.1) | <0.001 |
| Baseline R<sub>0</sub> (first R<sub>t</sub>) | 2.6 (1.7, 4.3) | 2.8 (1.9, 4.6) | 0.160 |
| Mean stringency (follow-up) | 64.5 (59.9, 70.9) | 57.4 (51.9, 64.1) | <0.001 |
| Latitude | 37.9 (36.7, 40.1) | 38.2 (34.5, 41.7) | 0.600 |
| Mean temp Jan–Jul (°F) | 56.0 (51.5, 58.8) | 55.7 (49.3, 62.3) | 0.790 |
| Follow-up days | 41 (22, 64) | 106 (90, 114) | <0.001 |
| **Region** |  |  | 0.040 |
| Northeast | 5 (2.9%) | 210 (7.5%) |  |
| Midwest | 70 (40.9%) | 919 (33.0%) |  |
| South | 79 (46.2%) | 1,320 (47.4%) |  |
| West | 17 (9.9%) | 336 (12.1%) |  |
| **Month of risk-set entry** |  |  | <0.001 |
| March | 26 (15.2%) | 843 (30.3%) |  |
| April | 115 (67.3%) | 1,366 (49.0%) |  |
| May | 23 (13.5%) | 285 (10.2%) |  |
| June | 5 (2.9%) | 169 (6.1%) |  |
| July | 2 (1.2%) | 122 (4.4%) |  |

We estimated the baseline reproduction number at risk-set entry for all 2,956 counties, and no county entered the risk set below the epidemic threshold (minimum R_0_: 1.22). The distribution was right-skewed (median 2.76, IQR 1.86–4.55; log R_0_ median 1.01, IQR 0.62–1.52) with substantial between-county variation, indicating that the minimum-case criterion for R_t_ estimability did not homogenize transmission intensity at entry. Basic reproduction number increased monotonically across tertiles of log population density (median R_0_ 2.00, 2.65, and 4.28 from lowest to highest tertile; Spearman ρ = 0.51), so denser counties entered the risk set with substantially higher baseline transmission potential. R_0_ distribution was similar between counties achieving and not achieving sustained control (median 2.6, IQR 1.7–4.3 versus 2.8, IQR 1.9–4.6, respectively; p = 0.16). No event occurred within three days of risk-set entry, so we derived the event definition and R_0_ from non-overlapping observation windows.

### Time to event analysis

Kaplan–Meier cumulative incidence of sustained control diverged by population density tertile from approximately calendar day 40 onward (Figure 2). By the end of follow-up, about 11%, 8%, and 3% of low-, medium-, and high-density counties, respectively, had achieved sustained control, with the high-density tertile lowest throughout.

**Figure 2:**
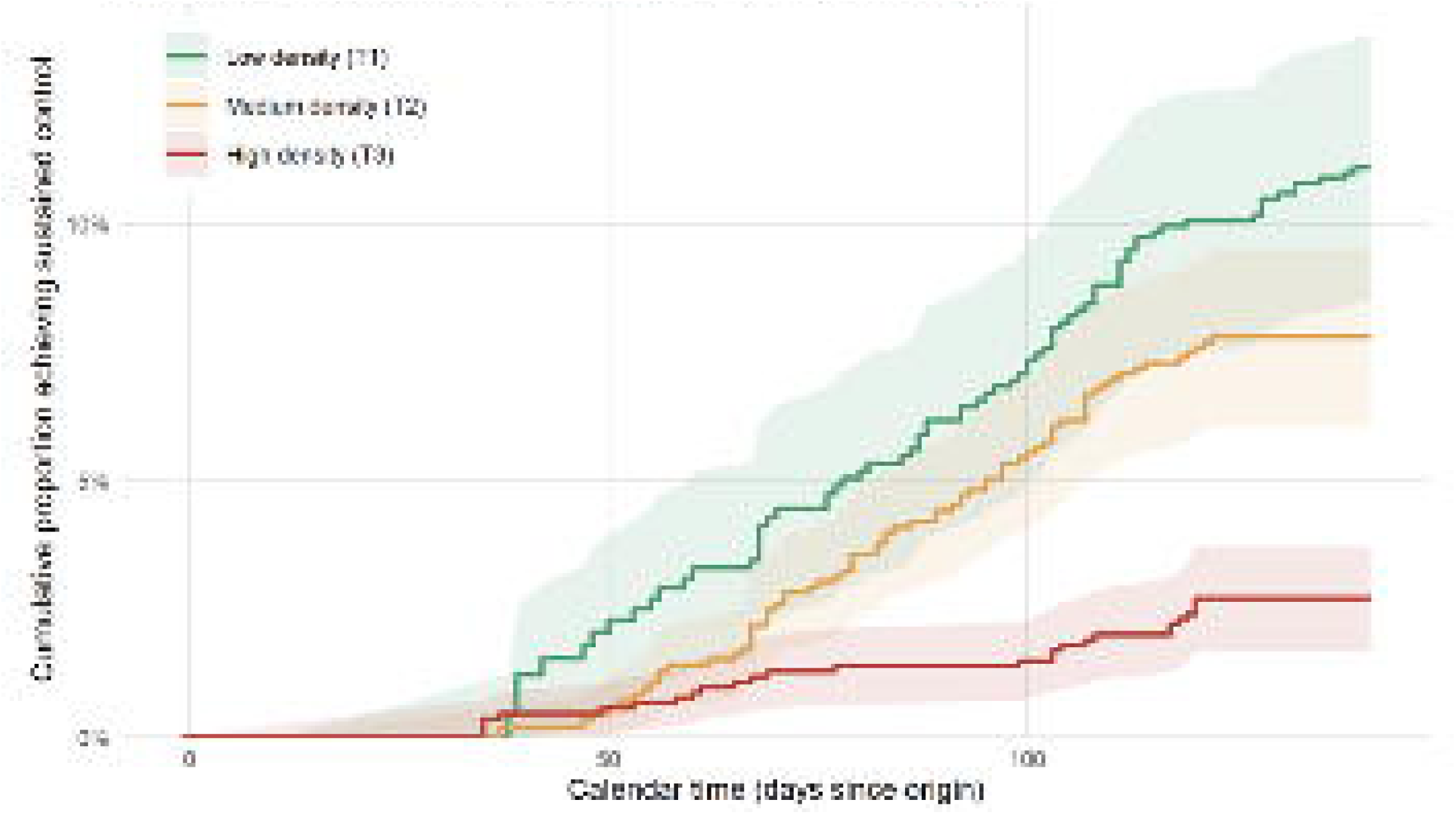
Cumulative incidence of sustained transmission control (mean R_t_*<* 1 for *>*3 consecutive days) over calendar time, stratified by tertile of county log-population-density. They-axis shows the cumulative proportion of counties having achieved the event, calculated as the complement of the Kaplan-Meier estimator with left-truncated entry. Overall, 5.8% of counties achieved the event.

In the multivariable mixed-effects Cox model, population density, stringency, and temperature were associated with the rate of sustained control, whereas R_0_ was not (Table 2). Each one-standard-deviation increase in log population density was associated with a 46% lower rate of achieving sustained control (HR 0.54, 95% CI 0.44–0.67). Higher stringency was associated with a faster rate of control (HR 1.50, 95% CI 1.09–2.05), which, consistent with the estimand, we interpret as an association conditional on state frailty. Temperature (HR 1.52, 95% CI 0.99–2.33), and R_0_ (HR per SD of log R_0_ 1.06, 95% CI 0.88–1.26) showed no statistically significant association.

**Table 2.**
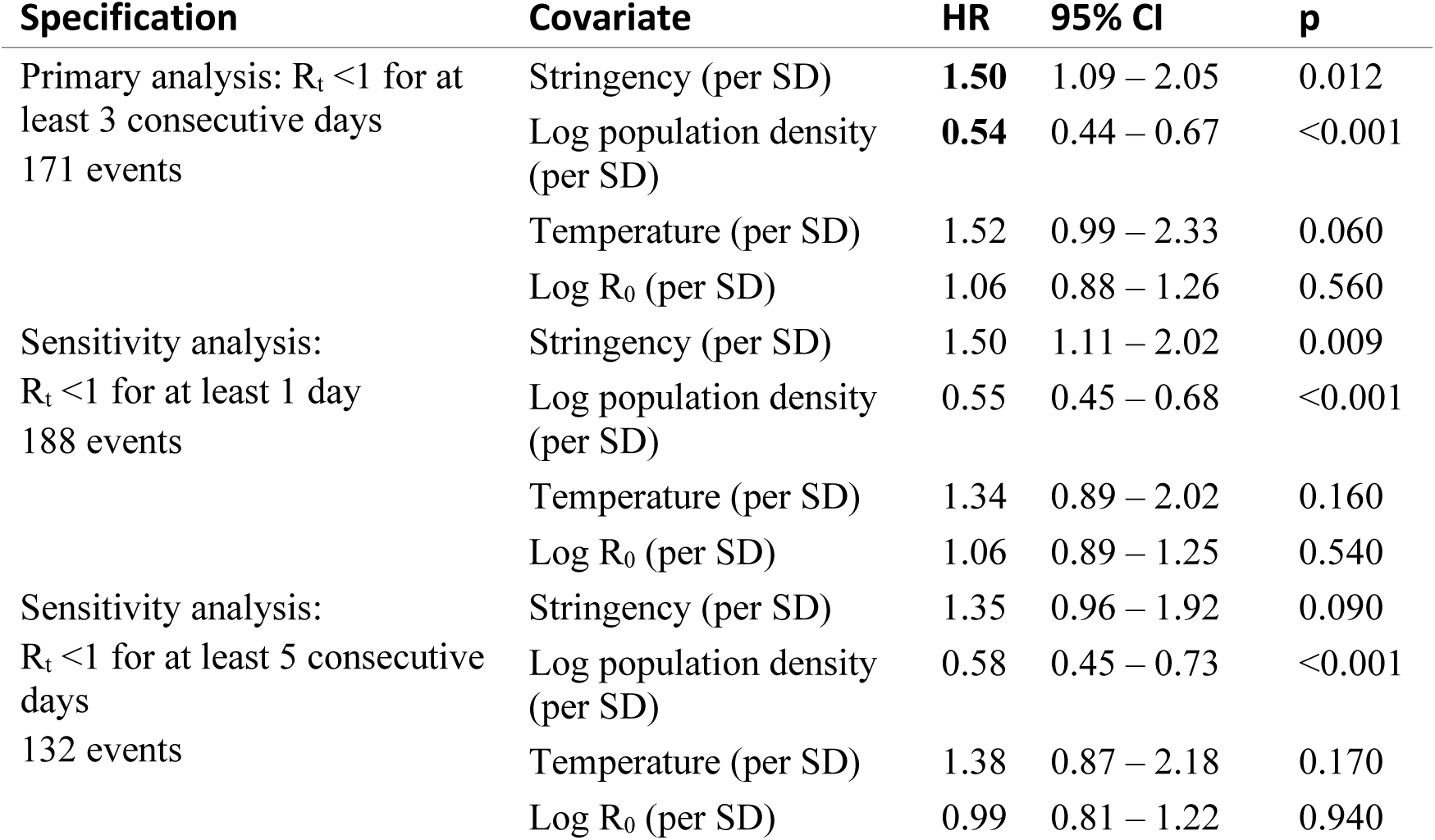
Mixed-effects Cox proportional-hazards model of time to sustained transmission control across alternative event definitions. Hazard ratios are per one-standard-deviation increase in each (scaled) covariate. All specifications include state-level Gaussian frailty. Frailty standard deviation on the log-hazard scale: 1.04 (primary), 1.18 (k = 1), 1.21 (k = 5). HR = hazard ratio.

The standard deviation of the state-level log-frailties was 1.04 on the log-hazard scale (variance 1.09) (Figure 3), indicating residual between-state heterogeneity beyond the four measured covariates: a state located one standard deviation above the national mean had about 2.8-fold the rate of control compared to an average state. This heterogeneity was comparable in magnitude to the measured fixed effects, though estimated from a sparse event distribution (Supplementary Table S1).

**Figure 3:**
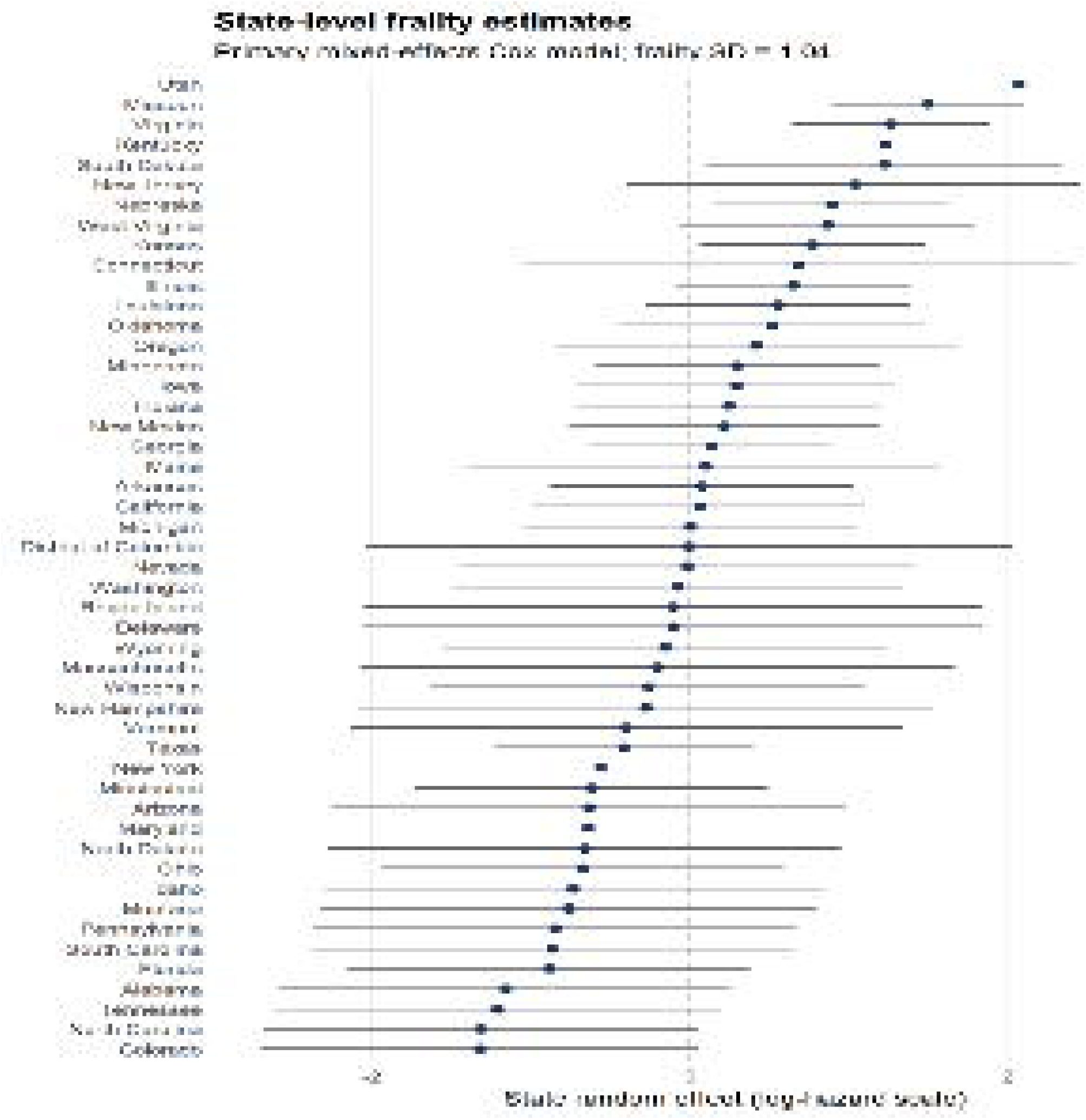
Forest plot of state-level random-effect (frailty) estimates from the primary mixed-effects Cox model, ordered by point estimate. Each row represents one state; horizontal bars show 95% prediction intervals on the log-hazard scale. The vertical reference line at zero indicates the population-m1erage state. The substantial spread (frailty SD *=* 1.04) indicates large unexplained heterogeneity in the speed of sustained control across states beyond what population density, stringency, temperature, and the baseline reproduction number capture.

### Diagnostics

The proportional-hazards assumption was supported globally (scaled Schoenfeld global test, χ²₄ = 4.78, p = 0.31) and individually for population density (p = 0.51), temperature (p = 0.76), and R_0_ (p = 0.38). Stringency showed a modest departure (χ²₁ = 4.07, p = 0.044), which the smoothed Schoenfeld residuals localized to the earliest weeks of follow-up, when the risk set was small and state policy responses were triggered directly by acute local outbreaks (see Discussion).

Restricting follow-up to calendar day 60 onward (140 of 171 events retained) removed the departure for stringency and globally, with all fixed-effect hazard ratios materially unchanged (Supplementary Table S2), though temperature itself showed a marginal departure within this window (χ²₁ = 3.63, p = 0.057). Reported hazard ratios are interpreted as time-averaged associations over the full study window.

Functional-form linearity was supported for all four covariates by likelihood-ratio tests against natural cubic splines, fitted one covariate at a time against the linear reference (population density p = 0.76; stringency p = 0.27; temperature p = 0.05; log R_0_ p = 0.17) and jointly (χ²₄ = 6.61, p = 0.16). A complementary polynomial-deviation test indicated departure for population density (χ²₂ = 10.9, p = 0.004) and, more weakly, temperature (χ²₂ = 6.36, p = 0.042). Because natural cubic splines are constrained to linearity beyond the boundary knots whereas polynomial terms are driven by the tails, this discrepancy reflects behaviour above the 95th percentile of population density; dfbeta localized it to a few high-density counties, none of which approached the Belsley influence threshold (Supplementary Material). Deviance residuals identified 0.06% of person-time intervals as exceeding |2.5|, consistent with the expected residual structure in counting-process Cox models. Discrimination was moderate (Harrell’s C = 0.66, 95% CI 0.63–0.70 from a 500-resample county-level cluster bootstrap).

### Sensitivity analyses

Table 2 summarizes hazard ratios under alternative event definitions of sustained control. Across all event definitions, the inverse association for log population density remained stable (HR 0.54– 0.58). Estimates for stringency were similar under the one-day (HR 1.50, 95% CI 1.11–2.02) and three-day primary (HR 1.50, 95% CI 1.09–2.05) definitions; under the five-day definition, the association attenuated (HR 1.35, 95% CI 0.96–1.92), reflecting both fewer events (132 vs 171) and reduced stringency variation among counties achieving prolonged control. The temperature association was positive but imprecise throughout. R_0_ consistently showed no association with time to control (HR 1.06, 1.06, and 0.99 for 1-, 3-, and 5-day definitions, respectively; Table 2). Entering R_0_ on its natural scale (rather than logarithmic) left conclusions unchanged (HR per SD of R_0_: 1.12, 95% CI 0.94–1.34; population density HR 0.53, 95% CI 0.43–0.66). Allowing the log R_0_ coefficient to vary before and after calendar day 60 demonstrated consistent period-specific hazard ratios (0.95 vs. 1.09, p = 0.58).

A quantitative bias analysis assessed whether differential expansion of testing (and, thus, of case ascertainment) across population densities could account for the observed population density association with the event under study (Figure 4). Adjusted R_t_ were reconstructed analytically and the model was refitted across assumed population-density differentials in testing expansion, ranging from no differential (γ=0) to 60% per population-density tertile. At zero differential the reconstruction reproduced the primary estimate exactly, as expected. Across the grid, the adjusted hazard ratio for population density remained stable (between 0.50 and 0.57).

**Figure 4:**
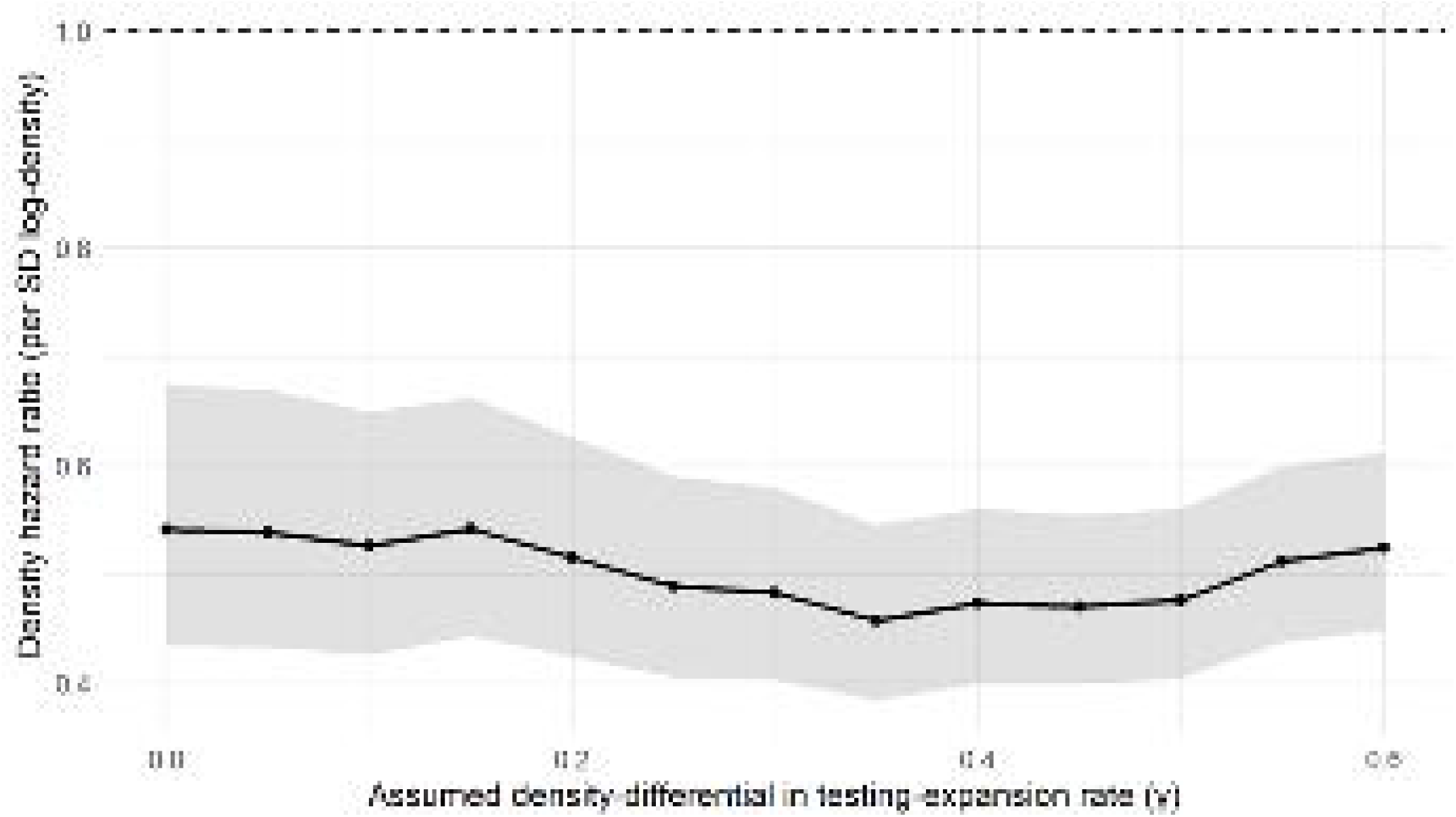
Sensitivity of the population-density hazard ratioto differential case ascertainment. Each point is the population-density hazard ratio (per SD of log population density) from the mixed-effects Cox model refit after reconstructing ascertainment-adjusted reproduction numbers under an assumed population-density differential in the rate of testing expansion (γ), ranging from no differential to a 60% difference per population-density tertile; shading is the 95% compatibility interval. *γ -*0 corresponds to no differential and reproduces the primary analysis exactly (HR 0.54). The dashed line marks the null. The hazard ratio remains well below 1 across the fall range, indicating that the population density association is not attributable to differential ascertainment operating through measured incidence; the γ axis is expressed in assumed units (.Methods).

Correct handling of estimability-induced left truncation was necessary for an unbiased population density estimate: ignoring delayed entry attenuated the log population density hazard ratio toward the null, whereas a gap-time clock biased it away from the null, so no naive simplification was conservative (Supplementary Figure S1).

## Discussion

In this analysis of 2,956 US counties during the first COVID-19 pandemic wave, sustained reductions in SARS-CoV-2 transmission below the epidemic threshold (R_t_<1) were rare (5.8% of counties) and concentrated among smaller, lower-density, predominantly Midwestern counties. Higher population density roughly halved the hazard of achieving sustained control per standard deviation of log population density, robustly across event definitions, model specifications, and diagnostic checks. County-specific R_0_ was not associated with the rate of control, and adjusting for it did not alter the association with population density with the study event: how far above the epidemic threshold a county began did not measurably determine how quickly it fell below it. Higher state-level policy stringency was associated with a moderately faster rate of control, though the estimate attenuated and became imprecise under the most conservative event definition. The higher crude mean stringency among counties achieving control partly reflects calendar-time exposure in the survival design and is addressed in the multivariable model through time-varying covariates. The temperature signal was directionally consistent with hypothesized seasonality but imprecise: with follow-up confined to a single wave (January–July 2020), the seasonal component of temperature cannot be separated from calendar time, leaving only spatial contrasts to inform the estimate. Substantial state-level residual heterogeneity indicated that factors beyond the measured covariates contributed meaningfully to variation in time to control.

Previous county-level studies in the United States have examined population density, social distancing and policy measures, and temperature as correlates of the level of the time-varying reproduction number (2–5), and non-pharmaceutical interventions have been linked to reductions in transmission at the national level (14). Our analysis differs in its outcome: rather than modeling the level of R_t_, it models the rate at which counties reach and sustain R_t_<1, a decision-relevant operational target. The two approaches are therefore complementary, and the persistence of a strong population-density gradient under a failure-time estimand that explicitly handles outcome-dependent observability strengthens the evidence that population density is a key structural correlate of early transmission control.

This framework carries practical consequences. Because a county’s entry into the risk set depends on meeting estimability thresholds, a process tied to population density, ignoring delayed entry misattributes unobserved event-free calendar time to low-density counties and biases the population density association toward the null. Conversely, using a gap-time clock discards the calendar-time baseline and biases it away from the null. The survival formulation resolves this by enforcing proper left truncation, while our quantitative bias analysis extends this logic to differential case ascertainment. Because testing capacity expanded unevenly during the first wave, differential ascertainment by population density is a plausible alternative explanation for the observed association, and it lies on a path that covariate adjustment cannot close. Explicitly quantifying this bias, rather than treating reported cases as complete, is a strength of this analysis: the population density hazard ratio remained between 0.50 and 0.57 under assumed differentials in testing expansion of up to 60% per population-density tertile. Calibrating this growth differential parameter against empirical, county-level testing surveillance data represents a logical future extension. The framework therefore transfers established failure-time methodology to a setting whose defining feature, that is, the outcome-dependent observability, has not been handled in prior county-level R_t_ analyses.

Several limitations need consideration. First, stringency exhibited a modest departure from the proportional-hazards assumption, during the earliest weeks of follow-up. We attribute this to reverse causation: states that moved most aggressively in mid-March did so in direct response to acute outbreaks, making early stringency partly endogenous to unobserved transmission intensity. Restricting follow-up to calendar day 60 onward recovers proportional hazards at minimal cost to substantive conclusions and is reported as supporting evidence rather than as a primary specification. Second, we held the serial-interval distribution constant at parameters appropriate for the ancestral strain, and county-level aggregation obscures sub-county heterogeneity in transmission and exposure, such as workplace settings, congregate facilities, and minority neighborhoods (19). Conversely, we preferred the county over the state as the unit of analysis because it captures the substantial within-state heterogeneity in transmission and yields more detailed, informative comparisons across a much larger number of units, at the cost of less precise Rt estimates in less populous counties. Third, the temperature association warrants particular caution: its seasonal component is collinear with calendar time and absorbed by the baseline hazard, so identification rests on spatial contrasts within calendar months, which are limited to a single wave (January–July 2020) and confounded with fixed county characteristics that the covariates and state frailty capture only partially. Fourth, the event definition treats the first sustained crossing as absorbing and does not address recurrent crossings; an Andersen–Gill recurrent-events extension(20) would be a natural next step for multi-wave data. Fifth, stringency was measured at the state level and assigned uniformly to all counties within a state, so it does not capture county-level policies or spillover from neighboring counties, including those across state borders. Sixth, both the event and the baseline reproduction number were defined from the posterior mean Rt, without accounting for its posterior uncertainty; a stricter definition requiring the upper credible limit of Rt to fall below one would have been preferable in principle, but yielded too few events in the first wave to support stable estimation. Finally, counties that brought transmission below the threshold while accruing very few cases never became estimable and contributed neither an entry nor an event. Because such counties are disproportionately small and low-density, this selection most plausibly removes early-control events from the low-density stratum specifically, attenuating rather than inflating the population density association; the reported association of lower population density with control is therefore likely conservative.

Several findings have implications for emerging-pathogen surveillance and response planning. The rarity of sustained control during the first wave (5.8% of counties) and its concentration in lower-density, smaller counties indicate that structural population characteristics were strongly associated with where early transmission control was achieved, with policy variation as a secondary modifier. The substantial state-level residual heterogeneity suggests that modifiable state-level factors, such as governance quality, healthcare capacity, mobility patterns, and variability in policy enforcement, are productive targets for future investigation. The survival framework offers a practical analytic tool for such work, complementing rather than replacing regression-on-R_t_ approaches. To our knowledge, this is the first analysis to apply this failure-time methodology, with estimability-induced left truncation, to time to sustained control of the reproduction number, and there is considerable room for further models to be developed and compared, including recurrent-event, multi-state, and competing-risk formulations, as well as joint models of transmission and case ascertainment. A further strength is that the framework is generic with respect to the event: the same left-truncated formulation can accommodate other operational targets, such as reaching a given percentage of intensive care unit capacity, and several such events can be analyzed simultaneously. Evaluating jurisdictions by their rate of reaching an operational target (R_t_<1) -rather than by average transmission level - provides a clearer, decision-relevant metric for prospective allocation decisions and public health interventions.

## Data Availability

All data produced in the present study are available upon reasonable request to the authors.

https://zenodo.org/records/21355447

---

**Supplementary Figure S1:**
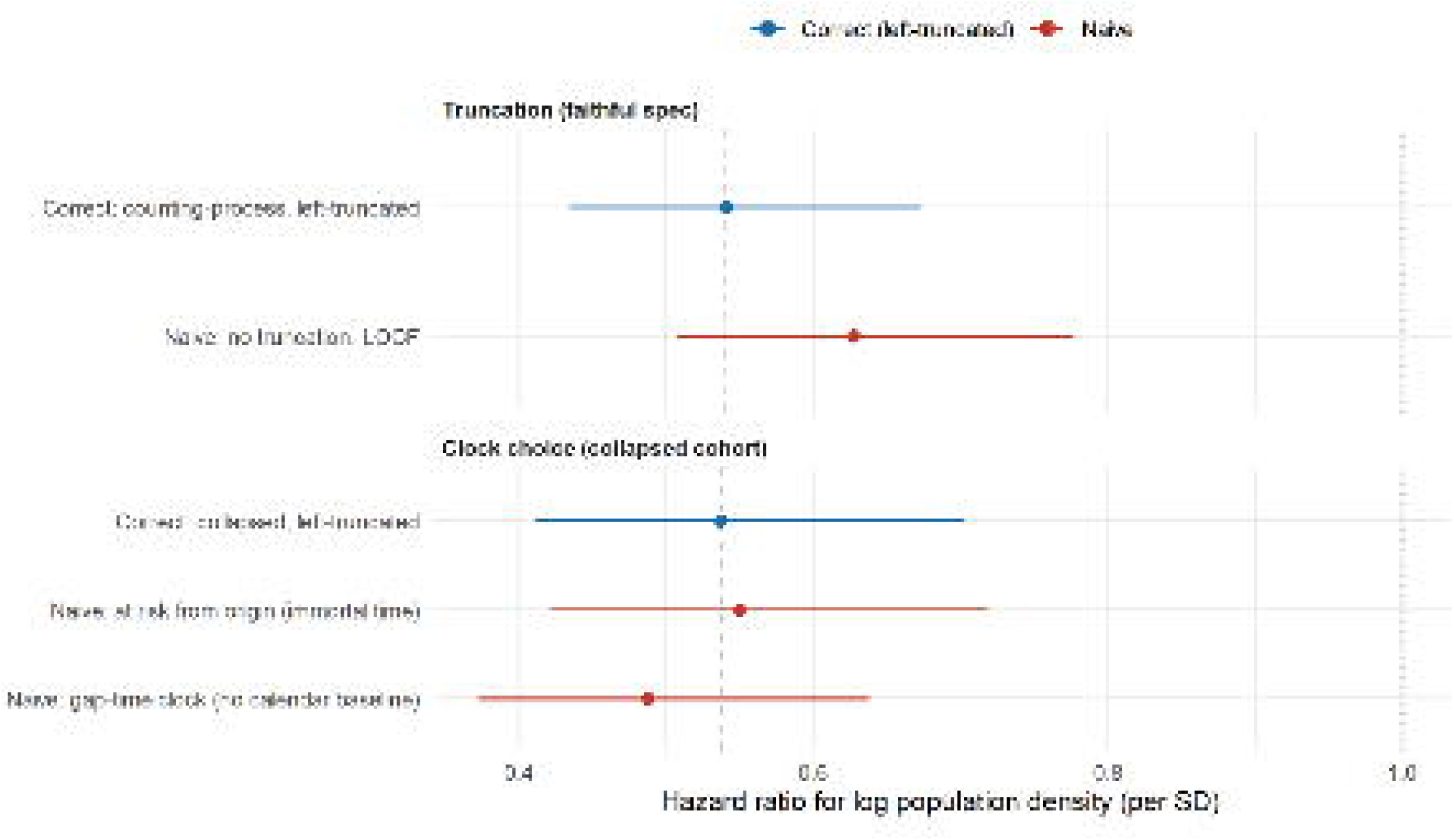
Mishandling estimability-induced left truncation biases the population-density hazard ratio. Points are hazard ratios for log population density (per standard deviation) from the mixed-effects Cox model; horizontal bars are 95% confidence intervals. In each panel, the dashed vertical line marks the correct (left-truncated) estimate and the dotted line marks the null (hazard ratio = 1). Upper panel, ‘Truncation (counting-process layout)’: compared with the fit accounting for left truncation, ignoring delayed entry attenuates the hazard ratio toward the null. Lower panel, ‘Clock choice (collapsed cohort)’: an at-risk-from-origin (immortal-time) clock also biases the hazard ratio toward the null, whereas a gaptime clock that discards the calendar-time baseline biases it away from the null. Specifications are defined in Methods. No naive simplification is conservative.

