## Supplementary Table S1, Supplementary Table S2, Supplementary Material for "Time to sustained control of the effective reproduction number as a failure-time estimand: a county-level analysis of the first SARS-CoV-2 wave in the United States"

The 171 events were distributed unevenly across the 49 frailty levels (48 contiguous states and the District of Columbia; Alaska and Hawaii contributed no estimable counties): median 1 event per state (IQR 0–5, range 0–20). Seventeen states contributed no events, and nine contributed a single event, so a minority of higher-event states (Kentucky, Missouri, Virginia, and Kansas together account for 66 of 171 events) largely drives the state-level frailty variance. The frailty standard deviation is therefore best read as an approximate summary of residual between-state heterogeneity rather than a precisely identified parameter; individual state random effects, particularly for zero-event states whose predictions are determined almost entirely by shrinkage, warrant caution.

**Table S1.** Counties at risk and events achieving sustained control, by state (48 contiguous states and the District of Columbia). Event count orders states.

| State | Counties at risk | Events | Event % |
|---|---|---|---|
| Kentucky | 118 | 20 | 16.9 |
| Missouri | 109 | 18 | 16.5 |
| Virginia | 131 | 17 | 13.0 |
| Kansas | 94 | 11 | 11.7 |
| Georgia | 159 | 9 | 5.7 |
| Illinois | 98 | 9 | 9.2 |
| Nebraska | 71 | 9 | 12.7 |
| Texas | 244 | 8 | 3.3 |
| Louisiana | 64 | 8 | 12.5 |
| Minnesota | 84 | 5 | 6.0 |
| Oklahoma | 72 | 5 | 6.9 |
| South Dakota | 58 | 5 | 8.6 |
| West Virginia | 51 | 5 | 9.8 |
| New Mexico | 30 | 5 | 16.7 |
| Iowa | 99 | 4 | 4.0 |
| Indiana | 92 | 4 | 4.3 |
| Arkansas | 75 | 4 | 5.3 |
| Utah | 11 | 4 | 36.4 |
| Michigan | 81 | 3 | 3.7 |
| California | 55 | 3 | 5.5 |
| Mississippi | 82 | 2 | 2.4 |
| Oregon | 32 | 2 | 6.2 |
| New Jersey | 21 | 2 | 9.5 |

| | | | |
|---|---|---|---|
| Ohio | 88 | 1 | 1.1 |
| Wisconsin | 72 | 1 | 1.4 |
| Florida | 67 | 1 | 1.5 |
| New York | 62 | 1 | 1.6 |
| Washington | 38 | 1 | 2.6 |
| Wyoming | 22 | 1 | 4.5 |
| Maine | 16 | 1 | 6.2 |
| Nevada | 13 | 1 | 7.7 |
| Connecticut | 8 | 1 | 12.5 |
| North Carolina | 100 | 0 | 0.0 |
| Tennessee | 95 | 0 | 0.0 |
| Alabama | 67 | 0 | 0.0 |
| Pennsylvania | 67 | 0 | 0.0 |
| Colorado | 62 | 0 | 0.0 |
| South Carolina | 46 | 0 | 0.0 |
| North Dakota | 43 | 0 | 0.0 |
| Idaho | 39 | 0 | 0.0 |
| Montana | 36 | 0 | 0.0 |
| Maryland | 24 | 0 | 0.0 |
| Arizona | 15 | 0 | 0.0 |
| Vermont | 14 | 0 | 0.0 |
| Massachusetts | 12 | 0 | 0.0 |
| New Hampshire | 10 | 0 | 0.0 |
| Rhode Island | 5 | 0 | 0.0 |
| Delaware | 3 | 0 | 0.0 |
| District of Columbia | 1 | 0 | 0.0 |

**Table S2.** Fixed-effect hazard ratios from the proportional-hazards sensitivity analysis restricting follow-up to calendar day 60 onward (140 of 171 events retained). Hazard ratios are per one-standard-deviation increase in each scaled covariate; state-level Gaussian frailty included.

| Covariate | Hazard ratio | 95% CI | p |
|---|---|---|---|
| Stringency (per SD) | 1.50 | 1.03 – 2.20 | 0.0360 |
| Log population density (per SD) | 0.50 | 0.39 – 0.64 | <0.001 |
| Temperature (per SD) | 1.39 | 0.81 – 2.38 | 0.230 |
| Log $R_0$ (per SD) | 1.09 | 0.89 – 1.33 | 0.400 |

**Supplementary Text. Functional form of the population-density association.**

Two complementary tests of linearity gave different answers for population density: the likelihood-ratio test against a natural cubic spline supported linearity ($p = 0.76$), whereas a polynomial-deviation test using quadratic and cubic terms indicated departure ($\chi^2_2 = 10.9$, $p = 0.004$). The two tests address different questions. Natural cubic splines are constrained to be linear beyond their boundary knots, placed here at the 5th and 95th percentiles of the covariate distribution, so the spline test is insensitive to behaviour in the tails. Polynomial terms are unbounded and are correspondingly dominated by them. The discrepancy therefore localises the departure to densities above the 95th percentile rather than to the body of the distribution.

Influence diagnostics identify the counties responsible. The largest dfbeta values for the log population density coefficient were Hudson County, New Jersey (0.029), Falls Church, Virginia (0.026), and Fairfax City, Virginia (0.023). None approached the Belsley influence threshold of $2/\sqrt{\text{events}} = 0.153$, and no county exceeded that threshold on any coefficient. The tail departure therefore did not materially affect the estimated coefficient, and the linear-in-log-hazard specification was retained for the primary model. The population density hazard ratio is accordingly interpreted as an average contrast over the observed population density range rather than as a constant gradient at its extremes.
